# Listening Difficulties in Children with Normal Audiograms: Relation to Hearing and Cognition

**DOI:** 10.1101/2020.10.05.20205468

**Authors:** Lauren Petley, Lisa L. Hunter, Lina Motlagh Zadeh, Hannah J. Stewart, Nicholette T. Sloat, Audrey Perdew, Li Lin, David R. Moore

## Abstract

**Objectives:** Children presenting at audiology services with caregiver-reported listening difficulties often have normal audiograms. The appropriate approach for the further assessment and clinical management of these children is currently unclear. In this Sensitive Indicators of Childhood Listening Difficulties (SICLiD) study we assessed listening ability using a reliable and validated caregiver questionnaire (the ECLiPS) in a large (n = 146) and heterogeneous sample of 6-13 year-old children with normal audiograms. Scores on the ECLiPS were related to a multifaceted laboratory assessment of the children’s audiological, psycho- and physiological-acoustic and cognitive abilities. This report is an overview of the SICLiD study and focuses on the children’s behavioral performance. The overall goals of SICLiD were to understand the auditory and other neural mechanisms underlying childhood listening difficulties and to translate that understanding into clinical assessment and, ultimately, intervention.

**Design:** Cross-sectional behavioral assessment of children with ‘listening difficulties’ and an age-matched ‘typically developing’ control group. Caregivers completed the ECLiPS and the resulting Total standardized composite score formed the basis of further descriptive statistics, univariate and multivariate modeling of experimental data.

**Results:** All scores of the ECLiPS, the SCAN-3:C, a standardized clinical test suite for auditory processing, and the NIH Cognition Toolbox were significantly lower for children with listening difficulties than for their typically developing peers, using group comparisons via *t*-tests and Wilcoxon Rank Sum tests. A similar effect was observed on the LiSN-S test for speech sentence-in-noise intelligibility, but only reached significance for the Low Cue and High Cue conditions, and the Talker Advantage derived score. Stepwise regression to examine the factors contributing to the ECLiPS Total scaled score (pooled across groups) yielded a model that explained 42% of its variance based on the SCAN-3:C composite, LiSN-S Talker Advantage, and the NIH Toolbox Picture Vocabulary and Dimensional Change Card Sorting scores (F_4,95_ = 17.35, *p* < 0.001). High correlations were observed between many test scores including the ECLiPS, SCAN-3:C and NIH Toolbox composite measures. LiSN-S Advantage measures generally correlated weakly and non-significantly with non-LiSN-S measures. However, a significant interaction was found between extended high frequency threshold and LiSN-S Talker Advantage.

**Conclusions:** Children with listening difficulties but normal audiograms have problems with the cognitive processing of auditory and non-auditory stimuli that include both fluid and crystallized reasoning. Analysis of poor performance on the LiSN-S Talker Advantage measure identified subclinical hearing loss as a minor contributing factor to talker segregation. Beyond auditory tests, evaluations of children with complaints of listening difficulties should include standardized caregiver observations and consideration of broad cognitive abilities.

## Introduction

Listening is an active form of hearing that implies a contribution of attention and is the basis of human auditory communication. Most people have listening difficulties (LiD) at one time or another, while others have LiD continuously. Although these difficulties are often accompanied by audiometric hearing loss, a substantial proportion of both children (Hind *et al*., 2011) and adults (Parthasarathy *et al*., 2020) attending audiology clinics, presumably due to LiD, are found to have normal audiometric sensitivity. Nationally within the US it has recently been estimated that about 80 million adults have self-identified LiD, of whom half have hearing loss, while the other half have clinically normal hearing (Edwards, 2020).

Individuals with LiD commonly have difficulty understanding speech in challenging situations such as noise, but also have other symptoms that can include difficulty following instructions and understanding rapid or degraded speech (Jerger & Musiek, 2000). While these difficulties have collectively been called auditory processing disorder (APD) (American Academy of Audiology, 2010; ASHA, 1996), the symptoms described require not only auditory processing, but also speech decoding and the successful deployment of selective attention towards task-relevant stimuli (Moore *et al*., 2010, 2018).

Despite the prevalence of LiD without hearing loss, listening is poorly understood clinically. For example, much of the controversy surrounding the concept of APD centers on whether LiD in children is due to a ‘hearing’ or to a ‘listening’ problem (Moore, 2018). It is beyond the scope of this study to revisit that controversy. Nevertheless, because APD remains a widely used diagnosis, especially in pediatric audiology where all are agreed that children with LiD represent an unmet need, it is important to understand and to parse the cognitive, biological, perceptual and social/academic components of LiD.

Both childhood and older age are marked by changes in attention, with the maturation of brain attention networks persisting through adolescence (Rueda & Posner, 2013), and losses that may be specific to dual set maintenance later in life (Verhaeghen & Cerella, 2002). A host of other top-down cognitive (Davis & Johnsrude, 2007) and bottom-up auditory (B. C. J. Moore, 2012) abilities underlie speech intelligibility and, in almost all of these abilities that have been studied, children generally perform more poorly than adults (Moore *et al*., 2011; Rueda & Posner, 2013; Sanes & Woolley, 2011). Listening necessarily involves higher areas of brain function, beyond the CANS, for example the “posterior hot zone”, a broad region of the posterior parietal, occipital and temporal cortex that appears to be the common locus of activation during different modes of conscious experience (Koch, 2018; Koch *et al*., 2016). The primary auditory cortex, in contrast, does “not directly contribute to the content of auditory … experience” (Koch, 2018).

Listening among individual children occurs along a continuum, as evidenced by caregiver questionnaires, including the Children’s Communication Checklist (CCC-2; Bishop, 2006) and the Evaluation of Children’s Listening and Processing Skills (ECLiPS; Barry *et al*., 2015; Barry & Moore, 2014; Roebuck & Barry, 2018). For some children, LiD is problematic because it impairs communication to a degree that substantially delays their social, academic and personal development relative to their peers.

Although this study focuses on LiD without clinical hearing loss, recent large-scale studies have shown that children with even mild hearing loss (better ear pure tone average, PTA of 21-40 dB HL) have poorer language skills, leading to impaired learning (Ching *et al*., 2018; Moeller & Tomblin, 2015; Moore *et al*., 2020). Mild hearing loss may go undetected clinically (e.g., ‘pass’ at neonatal hearing screen), or may not be followed up because of a perceived lack of agreement on how to proceed. In addition, the current clinical definition of ‘normal hearing’ does not guarantee the absence of peripheral pathology. Several forms of subclinical (or ‘hidden’) hearing loss have been identified, including minimal or slight hearing loss (PTA = 15 – 20 dB HL; Bess *et al*., 1998; Moore *et al*., 2020), extended high frequency (EHF) hearing loss (Hunter *et al*., 1996, 2020; Motlagh Zadeh *et al*., 2019), auditory synaptopathy (Kujawa & Liberman, 2019), and reduced spectral or temporal resolution of cochlear origin (Oxenham & Bacon, 2003). Subclinical hearing loss affects speech recognition in challenging listening conditions. For example, recent evidence in children with clinically normal hearing taking ototoxic drugs showed a significant positive relationship between EHF hearing loss and poor speech in noise performance (Blankenship *et al*., 2021). Studies in normal hearing adults have found significant correlation between poorer EHF thresholds and poorer speech in noise perception (Motlagh Zadeh *et al*., 2019; Yeend *et al*., 2019). In the broader field of learning disabilities (e.g., developmental language disorder, reading disorder), it is often assumed that children with clinically ‘normal’ hearing have problems that must lie outside the auditory system. Together, these sources point to a potentially large number of cases where LiD may involve an unrecognized origin in the ear.

Impaired function of the CANS is another possible source of LiD. For example, reversibly impaired binaural interaction and spatial hearing have been found consequent on earlier otitis media (Moore *et al*., 1991; Pillsbury *et al*., 1991; Whitton & Polley, 2011) and ear canal atresia (Wilmington *et al*., 1994) in children. The impairment is considered to be ‘central’ because it depends on binaural interaction, which only occurs in the brain, and because it can remain after the peripheral pathology resolves, leaving the ears audiometrically and tympanometrically normal. However, studies in this field generally did not consider the possibility of persistent subclinical hearing loss.

Otitis media earlier in life is also thought to underlie reduced spatial unmasking, termed ‘spatial processing disorder’ (SPD; Cameron *et al*., 2014, 2015; Graydon *et al*., 2017), as indexed by the Listening in Spatialized Noise – Sentences (LiSN-S) test. Spatial unmasking is a well-documented phenomenon in which spatially separating a target sound from a masking or competing sound renders the target more intelligible. The neural mechanisms of SPD remain unclear, and studies reporting SPD have not ruled out persistent subclinical hearing loss.

Reduced cognitive function is another possible mechanism of LiD. For example, executive control is crucial for establishing sustained goal-directed attention and dynamically switching attention based on task demands (Petersen & Posner, 2012; Posner & Petersen, 1990). Working memory supports speech comprehension, particularly under difficult listening conditions (Rudner & Rönnberg, 2008), and is associated with performance on even simple, non-speech hearing tests, such as frequency discrimination (Banai & Ahissar, 2004). Thus, poorer performance on those tasks may reflect either or both sensory and cognitive impairment (Moore *et al*., 2010; Sharma *et al*., 2014).

The primary purpose of the Sensitive Indicators of Childhood Listening Difficulties (SICLiD) study, a large, US National Institutes of Health funded, longitudinal research project, of which this is one of the first experimental reports, is to advance the understanding of APD, LiD and sub-clinical hearing loss to provide a basis for improvements in the diagnosis and management of children with LiD. Here, we present baseline behavioral data from the first full analysis of a large group of children with mixed listening abilities, as reported by their caregivers on the well-validated Evaluation of Children’s Listening and Processing Skills (ECLiPS) questionnaire (Barry *et al*., 2015; Barry & Moore, 2014; Roebuck & Barry, 2018). The primary aim of this study was to examine the relationship between hearing, listening and cognitive performance through univariate group comparisons and multivariate statistical modeling using quantitative metrics from a battery of auditory and cognitive tests. Based on previous research, we predicted that these analyses would demonstrate broad differences between LiD and TD groups with respect to cognitive, ‘auditory processing’, and speech-in-noise factors, and that all of these factors would be represented in the regression model predicting listening ability.

## Materials and Methods

This study was approved by the Institutional Review Board of Cincinnati Children’s Hospital (CCH) Research Foundation.

### Participants

Requirements for eligibility in the LiD group included caregiver-reported listening difficulties (see Procedure), age between 6 and 13 years old upon enrolment, English native language, and the absence of any neurologic, psychiatric or intellectual (IQ < 80) condition that would prevent or restrict ability to complete testing procedures. Typically-developing (TD) participants were eligible based on the same criteria, with the addition that they could not have listening difficulties or any ‘cognitive diagnosis’ (caregiver-reported developmental delay, attention, or learning disorder). Eligibility was determined based on caregiver responses on a medical and educational history ‘Background’ questionnaire.

A total of 166 participants (74 with LiD, 92 TD) was enrolled; 20 withdrew or otherwise exited the study (7 LiD, 13 TD). The remaining 146 participants ranged from 6.0 to 13.7 years of age at enrolment (Table 1). Due to attrition and updates to testing procedures, the sample size for each assessment was variable (see Results). All participants had clinically normal hearing, bilaterally, defined as pure tone thresholds ≤ 20 dB at all octave frequencies between 0.25 – 8 kHz.

**Table 1:** Participant numbers and demographic variables in the typically developing (TD) and listening difficulty (LiD) groups. Age is expressed as mean (SD). All other values are frequencies. Frequencies of “other cognitive concerns” reflect the number of individuals who had one or more caregiver-reported cognitive diagnoses.

|  | **TD** | **LiD** |
| --- | --- | --- |
| ***Total N*** | 79 | 67 |
| ***Age*** | 9.42 (2.04) | 9.79 (1.96) |
| ***Maternal Education Level*** |  |  |
| *High school and under* | 1 | 12 |
| *Some college and above* | 78 | 55 |
| ***Gender*** |  |  |
| *Female* | 33 | 23 |
| *Male* | 46 | 44 |
| ***Race*** |  |  |
| *Non-Caucasian* | 10 | 13 |
| *Caucasian* | 66 | 50 |
| *Multiracial* | 3 | 4 |
| ***Other Cognitive Concerns*** | 3* | 51 |

\* Three children in the TD group had received some form of speech or language intervention at school.

### Procedure

Participants with LiD were recruited initially from a medical record review study of over 1,100 children assessed for APD at Cincinnati Children’s Hospital. Of those children, 179 were diagnosed by an audiologist as having an auditory processing “disorder” and 364 as having an auditory processing “weakness” (for further details of the criteria for these diagnoses, see Moore *et al*., 2018). Caregivers of children in either of those categories who fit our inclusion criteria were invited to participate in this study. Over time, recruitment efforts were expanded to include children both with and without LiD from the broader community via print, electronic, social and digital media at hospital locations and in the local and regional area (e.g., grocery stores). Flyers were posted in relevant hospital clinics (i.e., Audiology, Developmental and Behavioral Pediatrics, Speech/Language Pathology) and sent out via email to hospital employees and families interested in research.

Interested caregivers completed eligibility screening procedures for their child by phone, or by paper or electronic questionnaires. Electronic questionnaires were delivered via a secure, unique link to REDCap, an electronic survey administration and database platform. Those who opted-in completed (i) a consent form, (ii) the Background questionnaire, (iii) the ECLiPS (Barry & Moore, 2014), to confirm eligibility for the LiD or TD group, and (iv) the CCC-2. If the child was deemed eligible by study staff, the caregiver was invited to schedule an initial visit.

Prior to completion of any other study-related procedures, caregivers reviewed the informed consent form with a study staff member and discussed the purpose, procedures, risks and benefits, duration, and expectations involved in the study. Children aged 11 and above were also assented using a child-friendly version of the consent document, per institutional policy. All participants received financial compensation for their participation.

About 70% (n=104) of the study sample scheduled individual visits. Most participants completed at least three separate visits, approximately 3 hours each. The average time between visits for the full sample was 18.6 days, but as some study procedures were added several months to a year after the start of the study, some participants returned to complete the series of visits several months apart over a 2-year period. Consequently, all age-related measures were adjusted for age at the time of testing (resolution = 0.1 years) rather than at enrolment. Audiometric thresholds were re-checked if the participant returned more than six months following their most recent audiogram. To maintain engagement between visits, participants were sent handwritten birthday and holiday cards throughout their enrolment. Caregivers also received periodic email communications about updates on the study website and newsletters about study and lab activities. Caregivers were encouraged to access the website, which was built as a tool to provide additional information about the study including interim results, staff members, and testing procedures.

As an alternative to scheduling individual visits, participants were offered the opportunity to attend a “summer camp” session, and approximately 30% of participants (n = 44) completed initial (‘Wave 1’) testing in this way. Two-day camp sessions were offered throughout the summer over the 2-year period of data collection. Six children were invited to attend each camp session and six camps were held each year, to an annual total of 36 camp spots. On the first day of camp, caregivers completed the informed consent process and then left for the day and returned to pick up their child in the afternoon. Across the two camp days, participants spent a total of 8-10 hours completing study procedures and the remaining 5-7 hours playing board games, having lunch and snacks, watching movies, and doing arts and crafts with other campers and the study staff. Participants rotated between 5-6 testing stations (sound booths, quiet testing rooms) with study staff to complete all of the different procedures. Overall, participants reported that they enjoyed participating in the study via summer camp sessions and expressed interest in returning for future sessions. While these differences in testing procedures raise the possibility of reduced motivation or fatigue (e.g., Key *et al*., 2017) in children who participated in the summer camps, the breaks provided between study procedures were intended to minimize these effects. Systematic differences between the children who participated in summer camp and those who did not prevent any causal inference regarding the potential influence of such factors on the present results, therefore no comparison of camp vs. no camp participants was performed.

### Materials

#### Background questionnaire

This questionnaire asked about the participant’s medical history, parental education, and demographic information. Key variables were date of birth, language, race and ethnicity, maternal education, history of ear and hearing problems (including pressure equalization, PE, tube placement), diagnosis of, or treatment for, learning problems (attention disorders, developmental delays, speech-language disorders), neurological (e.g. history of head trauma) or psychiatric conditions, school interventions, and birth history (prematurity, NICU stay). Demographic variables including maternal education, age, gender, and race, obtained from this questionnaire, are summarized in Table 1 for the listening difficulty (LiD) group and their peers (TD).

#### Caregiver evaluation of children’s listening (ECLiPS)

The ECLiPS (Barry & Moore, 2014) profiles children’s listening and communication abilities. It contains 38 simple statements (items) describing behaviors commonly observed in children. Caregivers were asked to rate how much they agreed with each statement on a five-point scale ranging from strongly disagree to strongly agree. The ratings were averaged to derive scores, which were scaled by age, on five subscales (speech & auditory processing, SAP; environmental & auditory sensitivity, EAS; language/ literacy/ laterality, LLL; memory & attention, M&A, and pragmatic & social skills, PSS) each containing 6-9 distinct items. These scales are further collected under Language, Listening, Social, and Total aggregate (composite) scores. All scales and composites were standardized for a population mean of 10 (s.d. = 3) based on British data (Barry *et al*., 2015). All of the subscales of the ECLiPS have high test-retest reliability, with intra-class correlations (ICCs) above 0.8 (Barry & Moore, 2014). Construct validity has been demonstrated through convergence with other established tests that measure similar skills, including the Children’s Auditory Processing Performance Scale (CHAPS; Smoski *et al*., 1998), the CCC-2 (Bishop, 2006), and the Social Communication Questionnaire (SCQ; Rutter *et al*., 2003). A secondary aim of this study was to provide initial US normative data for the ECLiPS and to compare them with the British norms.

### Caregiver evaluation of children’s communication (CCC-2)

The CCC-2 (Bishop, 2006) asks caregivers to respond to 70 items relating to the child’s communication skills. Subscales include speech, syntax, semantics, coherence, initiation, scripted language, context, nonverbal communication, social relations, and interests. The items are combined to create two composite scores, a General Communication Composite (GCC) and the Social Interaction Difference Index (SIDI). The GCC is a scaled composite of the scores on 8 of the 10 subscales, excluding subscales for social relations and interests.

### Audiometry

All participants were screened for normal hearing at standard, octave intervals from 0.25 - 8 kHz bilaterally. All except 27 (5 LiD, 22 TD) participants additionally completed threshold audiometry at the EHFs of 10, 12.5, 14 and 16 kHz using an Equinox audiometer (Interacoustics, Inc.) and the Hughson-Westlake adaptive method (ASHA, 2005b). Early in data collection, insert earphones (E-A-R TONE GOLD 3A) were used for standard frequencies and Sennheiser HDA 300 circumaural headphones for EHFs. For most children (n = 114; Hunter *et al*., 2021), the Sennheiser headphones were used for all frequencies. Participants were tested alternately with each ear first. They were instructed to respond when they heard a tone by pressing a response button or raising one hand. Those with elevated thresholds (> 20 dB HL) in any of the standard frequencies were excluded and were referred for further clinical care as appropriate. Participants were re-tested if they returned > 6 months after previous testing.

#### Auditory Processing Disorder (SCAN-3:C)

Participants were assessed in a sound attenuated booth or quiet room using the SCAN-3:C (Keith, 2009), a standardized test of APD for children aged 5-12 years (Emanuel *et al*., 2011). Stimuli were presented via a laptop PC through Sennheiser HD 215 headphones. The four diagnostic subtests of the SCAN-3:C are low-pass filtered words (FW), auditory figure-ground (AFG+8), competing words – directed ear (CW-DE), and competing sentences (CS). For FW, participants are asked to repeat low-pass filtered, monosyllabic words in quiet. AFG+8 is a speech-in-noise test requiring repetition of unfiltered monosyllabic words against multi-talker speech (the ‘noise’) at a fixed +8 dB signal/noise ratio (SNR). In CW-DE, two different monosyllabic words are presented simultaneously, one in each ear (i.e., dichotically), and participants report them back either left or right ear first. Similarly, for CS, unrelated sentences are presented to the left and right ears but, in this case, participants repeat only the sentence heard in one, directed ear per trial. The test-retest reliability for each of these SCAN subtests ranges between 0.64 and 0.73, while the test-retest reliability of the composite score is 0.77 (Keith, 2009). To establish its validity, the tests of the SCAN were based on the auditory processing abilities outlined in the ASHA 2005 Technical Report on (Central) Auditory Processing Disorders (ASHA, 2005a). A complete SCAN battery results in age-scaled scores for each subtest as well as a standardized composite.

### Speech hearing in noise (LiSN-S)

The Listening in Spatialized Noise – Sentences (LiSN-S) test (Brown *et al*., 2010; Cameron & Dillon, 2007; Phonak/NAL, 2011) measures the ability to listen and repeat simple, spoken sentences in the presence of informational masking, developed using the same criteria as the BKB sentences (Bench *et al*., 1979). The LiSN-S (US Edition; Brown *et al*., 2010) was administered using a commercial CD played on a laptop (Phonak/NAL, 2011), a task-specific soundcard, and Sennheiser HD 215 headphones. In the LiSN-S, participants are asked to repeat a series of target sentences (‘T’), presented directly in front (0°; diotic), while ignoring two distracting speakers (‘D1’, ‘D2’). There are four listening conditions, in which the distractors change voice (different or same as target) and/or position (0° and 90° virtual positions relative to the listener using generic head-related transfer functions; Humanski & Butler, 1988). The test is adaptive; the level of the target speaker decreases or increases in SNR relative to the distracting speech if the listener responds correctly or incorrectly during up to 30 sentences in each condition. The 50% correct SNR is either the Low cue speech reception threshold (SRT; same voice, 0° relative to the listener) or the High cue SRT (different voice, 90° relative to the listener). The three derived scores of the LiSN-S are the Talker Advantage, Spatial Advantage, and Total Advantage, so-called because each is the difference between SRTs from two conditions. This subtraction process should, to some extent, separate auditory from cognitive influences (Moore & Dillon, 2018), as discussed below. Test-retest comparisons for the four listening conditions of the LiSN-S show significant improvement from the first test to the second, but no differences for the advantage scores (Cameron *et al*., 2011).

### Cognition (NIH Toolbox)

Each participant’s cognitive skills were assessed using the NIH Toolbox Cognition Battery (Weintraub *et al*., 2013). Participants completed testing online or via an iPad app, in accordance with current Toolbox recommendations, in a private sound-attenuated booth or quiet room. The Battery contains up to eight standardized cognitive instruments measuring different aspects of fluid or crystallized reasoning. The precise composition of the testing battery is dependent on user choice and participant age.

All participants in this study completed the Picture Vocabulary test (PVT), Flanker Inhibitory Control and Attention test (Flanker), Dimensional Change Card Sort test (DCCS), and Picture Sequence Memory test (PSMT). Each test produced an age-corrected standardized score and the scores of all four tests were combined to calculate a single Early Childhood Composite. The PVT is an adaptive test in which the participant is presented with an audio recording of a word and selects which of four pictures most closely matches the meaning of the word. In the Flanker, which tests inhibition/attention, the participant reports the direction of a central visual stimulus (left or right, fish or arrow) in a string of five similar, flanking stimuli that may be congruent (same direction as target) or incongruent (opposite direction). The DCCS tests cognitive flexibility (attention-switching). Target and test card stimuli vary along two dimensions, shape and color. Participants are asked to match test cards to the target card according to a specified dimension that varies for each trial. Both the PVT and DCCS score accuracy and reaction time. PSMT assesses episodic memory by presenting an increasing number of illustrated objects and activities, each with a corresponding audio-recorded descriptive phrase. Picture sequences vary in length from 6-18 pictures depending on age, and participants are scored on the cumulative number of adjacent pairs remembered correctly over two learning trials.

Additional tests from the NIH Toolbox Cognition Battery were administered to all children 8 years of age and older. For the Fluid Composite measure, we used the list sorting working memory (LSWM) and the pattern comparison processing speed (PCPS) tests, in addition to the DCCS, Flanker, and PSMT, as above. LSWM assesses working memory by asking participants to arrange objects presented visually and auditorily (food and animals) in order of size. PCPS requires participants to respond as quickly as possible to whether two visually presented cards are the same or different. For the Crystallized Composite measure, participants completed the Reading Recognition (RR) test in addition to the PVT. The RR requires participants to read aloud words and letters accurately. A final Toolbox measure that was not part of a composite score, the Rey Auditory Verbal Learning (AVL) test measures verbal episodic memory as a supplement to the visual PSM test. In pediatric samples, the tests of the NIH Toolbox Cognition Battery demonstrate high reliability with short testing intervals between 7 and 21 days, with intraclass correlation coefficients (ICCs) between 0.76 and 0.99 (Weintraub *et al*., 2013a, b), but recent evidence suggests that their reliability over longer test-retest intervals (1-2 years), is substantially lower, with ICCs between 0.24 and 0.85 (Taylor *et al*., 2020). The convergent validity of the NIH Toolbox Cognition Battery instruments has been assessed against a range of published tests used in clinical practice that measure similar cognitive capacities (Weintraub *et al*., 2013a). These assessments yielded convergent validity correlations ranging between 0.48 and 0.93, suggesting that these instruments index the desired constructs (Weintraub *et al*., 2013b).

### Analysis

The primary analysis was divided into three sequential parts. Part 1 examined subgroup differences between children with LiD who had a formal diagnosis of APD (Dx subgroup; Moore *et al*., 2018) and children with LiD but no formal diagnosis (noDx subgroup). The purpose of this analysis was to determine whether these two subgroups could be treated as a homogeneous LiD group for subsequent analyses. These subgroups were tested for differences on the ECLiPS Total scaled score, SCAN-3:C composite scaled score, LiSN-S Talker Advantage, Spatial Advantage, and Low Cue scores, and the NIH Cognition Toolbox Fluid, Crystallized, and Early Childhood Composites, as well as the AVL test. A potential influence of maternal education was first explored via a separate two-way ANOVA for each of these variables with the factors diagnosis (Dx, noDx) and maternal education level (collapsed into two groups: graduated high school or less, and some college or more). In no case was there a main effect of, or an interaction with, maternal education (*p* > 0.05). All group-level comparisons for the Dx versus noDx subgroups were therefore conducted using two-tailed, two-sample Student’s *t*-tests, when the assumption of normality was met, and Wilcoxon Rank-Sum tests in all other cases. To maximize sensitivity to any possible differences between these groups, and thus to allay any concerns that the groups might be combined inappropriately, the *p* values used for this analysis were not adjusted for multiple comparisons.

Part 2 of the analysis examined group differences between TD children and children with LiD (combined Dx subgroups) on all of the same measures used for Part 1. The LiD group was not combined across Dx subgroups for any test that demonstrated differences between the subgroups in Part 1 of the analysis. As in Part 1, a potential influence of maternal education was first explored via separate two-way ANOVAs, this time with the factors listening difficulties (TD, LiD) and maternal education level (levels as above). In no case was there a main effect of, or an interaction with, maternal education (*p* > 0.05). All group-level comparisons between LiD and TD groups were therefore assessed via two-tailed, two-sample Student’s *t*-tests. To control for inflation, *p* values were adjusted for multiple comparisons. Taking the position that each of the test instruments administered here targets auditory and/or cognitive capacities in a different way, each test (e.g., the LiSN-S, the NIH Cognition Toolbox) was treated as its own “family”. This was considered preferable to an alternative MANOVA approach within each test instrument due to the tendency for MANOVAs to yield ambiguous results for all dependent variables beyond that with the highest-priority when these variables are highly positively correlated (Tabachnick & Fidell, 2001). Thus, *p*-values were adjusted for the following number of comparisons: one comparison for the ECLiPS, two comparisons for the CCC-2, five comparisons for the SCAN-3:C, four comparisons for the LiSN-S, and thirteen comparisons for the NIH Cognition Toolbox. Similar adjustments were made for comparisons versus the normative sample on all standardized tests, except that the LiSN-S was adjusted for three comparisons since the “Pattern Scores” were not compared with the normative sample, and the NIH Cognition Toolbox scores were adjusted for eleven comparisons since the Dx and noDx LiD subgroups were combined for all of these tests.

Part 3 of the analysis aimed to identify the functional domains that contribute to LiD across all participants by using stepwise multiple regression to predict ECLiPS Total scaled scores. Given that the addition of more independent variables to a multiple regression increases the number of cases required to achieve the same statistical power (Tabachnick & Fidell, 2001), this analysis leveraged the continuous nature of listening skills (and consequently, ECLiPS scores) by combining data across the LiD and TD groups. Candidate predictors for this analysis included only assessments that were available across the full age range of the study sample. As discussed above, some tests of the NIH Cognition Toolbox did not meet this criterion. Candidate predictors included maternal education level, race, SCAN-3:C Composite Score, four LiSN-S scores (Low Cue, High Cue, Talker Advantage, Spatial Advantage), and four NIH Cognition Toolbox tests (DCCS, Flanker, PST, PVMT).

As previously discussed, EHF hearing has recently been identified as an important factor influencing speech comprehension in noise (Blankenship *et al*., 2021; Motlagh Zadeh *et al*., 2019; Yeend *et al*., 2019). To explore a possible role of EHF hearing thresholds, for which main effects were examined in a recent paper (Hunter *et al*., 2021), four interactions were included as candidate predictors. Three interactions with LiSN-S scores (Low Cue, Talker Advantage, Spatial Advantage) explored a possible contribution from EHF thresholds to speech-in-noise performance in complex environments. In light of the known benefit that linguistic proficiency provides under challenging listening conditions (Kaandorp *et al*., 2016), an interaction with the NIH PVT was also explored to examine the possible role of language skills in mitigating the influence of EHF hearing loss. In all four interactions, EHF hearing thresholds were computed as a single average across both ears and all frequencies (10, 12.5, 14, and 16 kHz).

All candidate predictors for the stepwise multiple regression (Part 3) were first examined against the ECLiPS Total scaled score via univariate analyses. Continuous candidate predictors were examined using Spearman’s Rho, while categorical predictors were examined via Student’s *t*-tests. Variables with *p* < 0.1 in the univariate analysis were included in the stepwise regression. Collinearity among these variables was examined using the Variance Inflation Factor, which ranged from 1.31 to 3.01, indicating no collinearity. Statistical comparison of participants with complete and incomplete data across all demographic variables and candidate predictors yielded no significant differences between the two groups. It was therefore assumed that data were missing at random. All data were included in the stepwise regression via the use of SAS v9.4 (The SAS Institute, Cary, NC) PROC MIXED with restricted maximum likelihood estimation. The stepwise selection process was conducted automatically by a SAS procedure with a series of alternating forward selection and backward elimination steps using a threshold of 0.1. Among all the candidate predictors, SCAN-3:C Composite Score, NIH PVT, LiSN Talker Advantage, NIH DCCS, and Maternal Education Level were selected to enter the model sequentially. To achieve a parsimony, the final model included the only four significant variables which explained 42% of the variance in ECLiPS Total Score.

Additional, secondary analyses examining correlations between factors other than the ECLiPS were used to follow up on results revealed by the primary analysis. Multiple Spearman’s Rho correlations were used in this analysis.

## Results

### Diagnosis of APD

A total of 20 participants with LiD had received a previous diagnosis of auditory processing disorder or weakness (Moore *et al*., 2018). Of the remaining 47 participants with LiD, 22 had audiological consultations, but received no diagnosis, and 25 had not sought audiological assessment. The results from univariate statistical comparisons between those with a diagnosis or weakness (Dx subgroup) and those with no diagnosis (noDx subgroup) are summarized in Table 2. Among children with LiD, the Dx subgroup did not differ significantly from the noDx subgroup on any measure listed in Table 2, with the exception of two cognitive verbal scores, the NIH Crystallized Composite and the AVL. In both cases, considered further below, children with a diagnosis of APD had significantly lower scores than those with no diagnosis. For the majority of the remaining analysis, all children with LiD were considered as a single group.

**Table 2:** Univariate comparisons of the LiD subgroups with (Dx) and without (noDx) an auditory processing disorder or weakness. Test statistics include W for Wilcoxon Rank-Sum tests and *t* values for *t*-tests. Reported *p*-values are not adjusted for multiple comparisons.

|  | noDx |  |  | Dx |  |  | Test Stat. | *p* | *d* |
| --- | --- | --- | --- | --- | --- | --- | --- | --- | --- |
|  | N | Mean | 95% CI | N | Mean | 95% CI |  |  |  |
| *ECLiPS Composite Scores* |  |  |  |  |  |  |  |  |  |
| *Total* | 47 | 3.00 | 2.54, 3.45 | 20 | 2.85 | 1.90, 3.80 | *t* = 0.32 | 0.753 | 0.08 |
| *SCAN-3:C* |  |  |  |  |  |  |  |  |  |
| *Comp.* | 43 | 95.12 | 91.65, 98.58 | 16 | 94.13 | 87.23, 101.02 | *t* = 0.31 | 0.784 | 0.08 |
| *LiSN-S Cue Scores* |  |  |  |  |  |  |  |  |  |
| *Low* | 44 | -0.64 | -1.02, -0.27 | 20 | -0.97 | -1.38, -0.55 | *t* = 1.02 | 0.313 | 0.27 |
| *LiSN-S Advantage Scores* |  |  |  |  |  |  |  |  |  |
| *Spatial* | 44 | -0.61 | -1.12, -0.10 | 20 | -0.75 | -1.49, -0.01 | *t* = 0.31 | 0.761 | 0.08 |
| *Talker* | 44 | -0.60 | -0.86, -0.34 | 20 | -0.42 | -0.95, 0.11 | *t* = -0.67 | 0.506 | 0.18 |
| *NIH Cognition Toolbox Tests* |  |  |  |  |  |  |  |  |  |
| *AVL* | 25 | -0.85 | -1.59, -0.12 | 16 | -2.47 | -3.52, -1.42 | *t* = -2.55 | 0.015 | 0.82 |
| *NIH Cognition Toolbox Composite Scores* |  |  |  |  |  |  |  |  |  |
| *Fluid* | 30 | 85.15 | 79.60, 90.70 | 15 | 84.50 | 74.85, 94.15 | *t* = 0.12 | 0.904 | 0.04 |
| *Cryst.* | 37 | 94.47 | 91.13, 97.81 | 17 | 87.99 | 80.76, 95.21 | W = 194.5 | 0.026 | 0.54 |
| *EC* | 34 | 90.83 | 86.50, 95.17 | 14 | 84.43 | 75.54, 93.32 | *t* = 1.42 | 0.162 | 0.45 |

### Effects of LiD

In univariate statistical comparisons, most measures showed significantly poorer performance for the LiD group than for the TD group (Table 3; Figs. 1-3). By design, ECLiPS Total scores were much lower [*t*(141.03) = 22.67, *p* < 0.001, *d* = 3.67] for children in the LiD group than for those in the TD group (Table 3 and Fig. 1). Note that two children in the LiD group had ECLiPS scores within the normal range on at least one subscale. These children were both in the Dx subgroup that had previously been diagnosed with APD. A repeated-measures ANOVA within the TD group demonstrated that the mean standardized Total and subscale scores did not differ significantly from each other [*F*(4, 312) = 1.18, *p* = 0.32, η^2^_G_ = 0.01], as would be expected if the UK and US samples were drawn from the same population. One-sample *t*-tests versus the normalization sample, however, suggest that the TD sample scored slightly, but significantly higher than the UK normative group on the Total score [*t*(78) = 2.98, *p* = 0.004, *d* = 0.34, mean = 10.81]. As expected, the LiD group scored considerably lower [*t*(66) = -32.73, *p* < 0.001, *d* = 4.00, mean = 2.96]. An ANOVA performed within the ECLiPS subscores for the LiD group showed a significant difference between the ECLiPS subscale scores, and this difference remained significant following Greenhouse-Geisser correction [*F*(3.12, 206.2) = 15.94, *p* < 0.001, η^2^_G_ = 0.11]. Note that the SAP subscale reflects the profile statements that are most closely aligned with listening. SAP ratings of children in the LiD group were all within the lowest 30^th^ percentile based on the test’s standard (UK) normalization, while at least one child in the LiD group scored in the top 25^th^ percentile on each of the other subscales. ECLiPS subscales and their relation to other measures will be considered further in a separate report.

**Figure 1:**
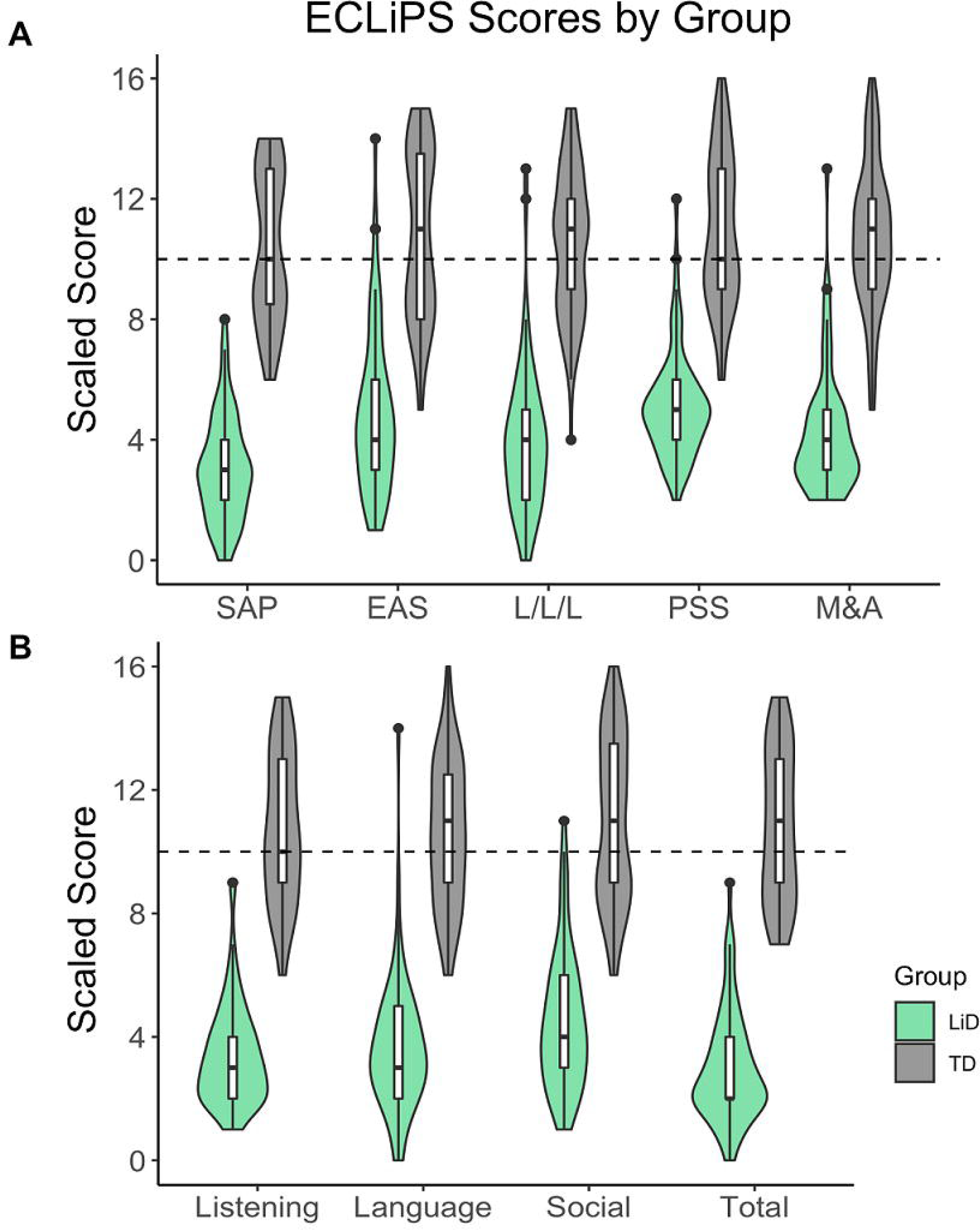
ECLiPS scores show better caregiver-assessed listening skills in the typically developing (TD) than in the listening difficulty (LiD) group of children. Violin plots with trimmed tails demonstrate the probability density of the data and are overlaid with boxplots indicating the median and interquartile range.Scores are shown for (A) all five ECLiPS subscales (speech & auditory processing, SAP; environmental & auditory sensitivity, EAS; language/ literacy/ laterality, LLL; memory & attention, M&A, and pragmatic & social skills, PSS) as well (B) as its four composite scores (Language, Listening, Social, and Total aggregate). Horizontal, dashed lines reflect expected standard score (here 10) for all scales in all figures.

**Table 3:** Univariate comparisons of the LiD and TD groups. Test statistics for group comparisons, when performed, include W for Wilcoxon Rank-Sum tests or *t* values for *t*-tests. All *p*-values are adjusted for multiple comparisons.

|  | TD |  |  | LiD |  |  |  |  |  |
| --- | --- | --- | --- | --- | --- | --- | --- | --- | --- |
|  | N | Mean | 95% CI | N | Mean | 95% CI | Test Stat. | *p* | *d* |
| ***ECLiPS Subtests*** |  |  |  |  |  |  |  |  |  |
| *SAP* | 79 | 10.39 | 9.85, 10.93 | 67 | 3.00 | 2.58, 3.42 |  |  |  |
| *EAS* | 79 | 10.78 | 10.17, 11.40 | 67 | 4.84 | 4.21, 5.47 |  |  |  |
| *L/L/L* | 79 | 10.52 | 9.99, 11.05 | 67 | 3.90 | 3.33, 4.47 |  |  |  |
| *M&A* | 79 | 10.72 | 10.19, 11.25 | 67 | 4.15 | 3.66, 4.63 |  |  |  |
| *PSS* | 79 | 10.92 | 10.38, 11.47 | 67 | 5.12 | 4.70, 5.54 |  |  |  |
| ***ECLiPS Composite Scores*** |  |  |  |  |  |  |  |  |  |
| *Listen.* | 79 | 10.73 | 10.21, 11.26 | 67 | 3.37 | 2.98, 3.77 |  |  |  |
| *Lang.* | 79 | 10.58 | 10.07, 11.09 | 67 | 3.61 | 3.10, 4.12 |  |  |  |
| *Social* | 79 | 11.00 | 10.42, 11.58 | 67 | 4.46 | 3.94, 4.99 |  |  |  |
| *Total* | 79 | 10.81 | 10.28, 11.34 | 67 | 2.96 | 2.53, 3.38 | *t* = 22.67 | < 0.001 | 3.67 |
| ***CCC-2 Composite Scores*** |  |  |  |  |  |  |  |  |  |
| *GCC* | 79 | 111.85 | 109.94, 113.76 | 64 | 81.70 | 78.33, 85.07 | *t* = -15.96 | < 0.001 | 2.68 |
| *SIDI* | 79 | -0.06 | -1.29, 1.16 | 64 | 2.27 | -0.57, 5.10 | *W* = 2871 | 0.32 | 0.27 |
| ***SCAN-3:C*** |  |  |  |  |  |  |  |  |  |
| *FW* | 69 | 13.00 | 12.54, 13.46 | 59 | 11.81 | 11.32, 12.30 | *W* = 1398 | 0.009 | 0.61 |
| *AFG* | 69 | 11.41 | 10.86, 11.95 | 59 | 9.59 | 8.92, 10.27 | *W* = 1243 | 0.001 | 0.73 |
| *CWDE* | 69 | 10.13 | 9.53, 10.73 | 59 | 7.00 | 6.31, 7.69 | *t* = -6.73 | < 0.001 | 1.19 |
| *CS* | 69 | 10.84 | 10.30, 11.38 | 59 | 8.71 | 7.96, 9.46 | *t* = -4.62 | < 0.001 | 0.82 |
| *Comp.* | 69 | 110.09 | 107.50, 112.67 | 59 | 94.85 | 91.73, 97.96 | *t* = 7.44 | < 0.001 | 1.32 |
| ***LiSN-S Cue Scores*** |  |  |  |  |  |  |  |  |  |
| *Low* | 76 | -0.13 | -0.35, 0.09 | 64 | -0.74 | -1.03, -0.45 | *t* = 3.32 | 0.004 | 0.56 |
| *High* | 76 | 0.28 | 0.05, 0.52 | 64 | -0.40 | -0.73, -0.08 |  |  |  |
| ***LiSN-S Advantage Scores*** |  |  |  |  |  |  |  |  |  |
| *Spatial* | 76 | -0.23 | -0.53, 0.08 | 64 | -0.65 | -1.07, -0.24 | *t* = -1.64 | 0.41 | 0.28 |
| *Talker* | 76 | -0.05 | -0.24, 0.13 | 64 | -0.54 | -0.79, -0.30 | *t* = 3.22 | 0.006 | 0.55 |
| *Total* | 76 | 0.40 | 0.16, 0.64 | 64 | -0.03 | -0.33, 0.28 |  |  |  |
| ***NIH Cognition Toolbox Tests*** |  |  |  |  |  |  |  |  |  |
| *FT* | 57 | 100.64 | 96.78, 104.49 | 57 | 89.40 | 85.59, 93.20 | *t* = -4.07 | 0.001 | 0.76 |
| *DCCS* | 57 | 104.11 | 100.25, 107.96 | 57 | 88.17 | 84.68, 91.66 | *t* = -6.01 | < 0.001 | 1.13 |
| *LSWM* | 67 | 109.77 | 106.09, 113.46 | 56 | 92.80 | 89.11, 96.50 | *t* = -6.32 | < 0.001 | 1.14 |
| *PCPS* | 52 | 103.29 | 97.27, 109.31 | 54 | 84.18 | 77.98, 90.39 | *t* = -4.33 | < 0.001 | 0.84 |
| *PSMT* | 73 | 110.90 | 106.32, 115.48 | 52 | 94.51 | 89.10, 99.93 | *t* = 4.53 | < 0.001 | 0.82 |
| *PVT* | 72 | 112.88 | 109.41, 116.35 | 59 | 97.36 | 94.04, 100.68 | *t* = -6.24 | < 0.001 | 1.10 |
| *RR* | 64 | 106.85 | 103.76, 109.94 | 55 | 89.91 | 86.77, 93.05 | *t* = -7.50 | < 0.001 | 1.38 |
| *AVL* | 48 | 0.66 | 0.22, 1.09 | 41 | -1.48 | -2.13, -0.84 |  |  |  |
| ***NIH Cognition Toolbox Composite Scores*** |  |  |  |  |  |  |  |  |  |
| *Fluid* | 51 | 109.13 | 104.28, 113.97 | 45 | 84.93 | 80.10, 89.77 | *t* = 7.64 | < 0.001 | 1.41 |
| *Cryst.* | 63 | 110.71 | 107.44, 113.97 | 54 | 92.43 | 89.14, 95.71 |  |  |  |
| *Total* | 50 | 112.26 | 108.13, 116.40 | 44 | 86.56 | 82.40, 90.72 |  |  |  |
| *EC* | 56 | 111.94 | 107.74, 116.15 | 48 | 88.97 | 84.91, 93.02 | *t* = 7.64 | < 0.001 | 1.50 |

The GCC composite score of the CCC-2 was significantly lower in the LiD than in the TD group [*t*(141) = -15.96, *p* < 0.001, *d* = 2.68], as shown in Table 3, and correlated highly with the ECLiPS total score [r*_s_*(143) = 0.75, *p* < 0.001]. Both groups were significantly different from the CCC-2 normative sample on the GCC composite score, with the TD group scoring higher than the sample [*t*(78) = 12.14, *p* < 0.001, *d* = 1.37, mean = 111.85] and the LiD group scoring lower [*t*(63) = -10.64, *p* < 0.001, *d* = 1.33, mean = 81.70]. The SIDI did not differ significantly between groups [W = 2871, *p* = 0.32, *d* = 0.27] and was uncorrelated with the ECLiPS [r*_s_*(143) = -0.060, *p* = 0.477].

SCAN-3:C composite scores were lower [*t*(126) = 7.44, *p* < 0.001, *d* = 1.32] for the LiD group than for the TD group. This difference can be seen in Fig. 2 and Table 3. Follow-up *t*-tests revealed significant differences between the LiD group and their TD peers on each of the SCAN-3:C subtests (details in Table 3). Results obtained on the composite measure thus reflect all functional domains tested by the SCAN-3:C. Similarly to other tests, both the TD and LiD group SCAN-3:C composite scores were significantly different from the normative sample, with the TD group scoring slightly higher [*t*(68) = 7.64, *p* < 0.001, *d* = 0.92, mean = 110.09] and the LiD group scoring slightly lower [*t*(58) = -3.24, *p* = 0.002, *d* = -0.42, mean = 94.85].

**Figure 2:**
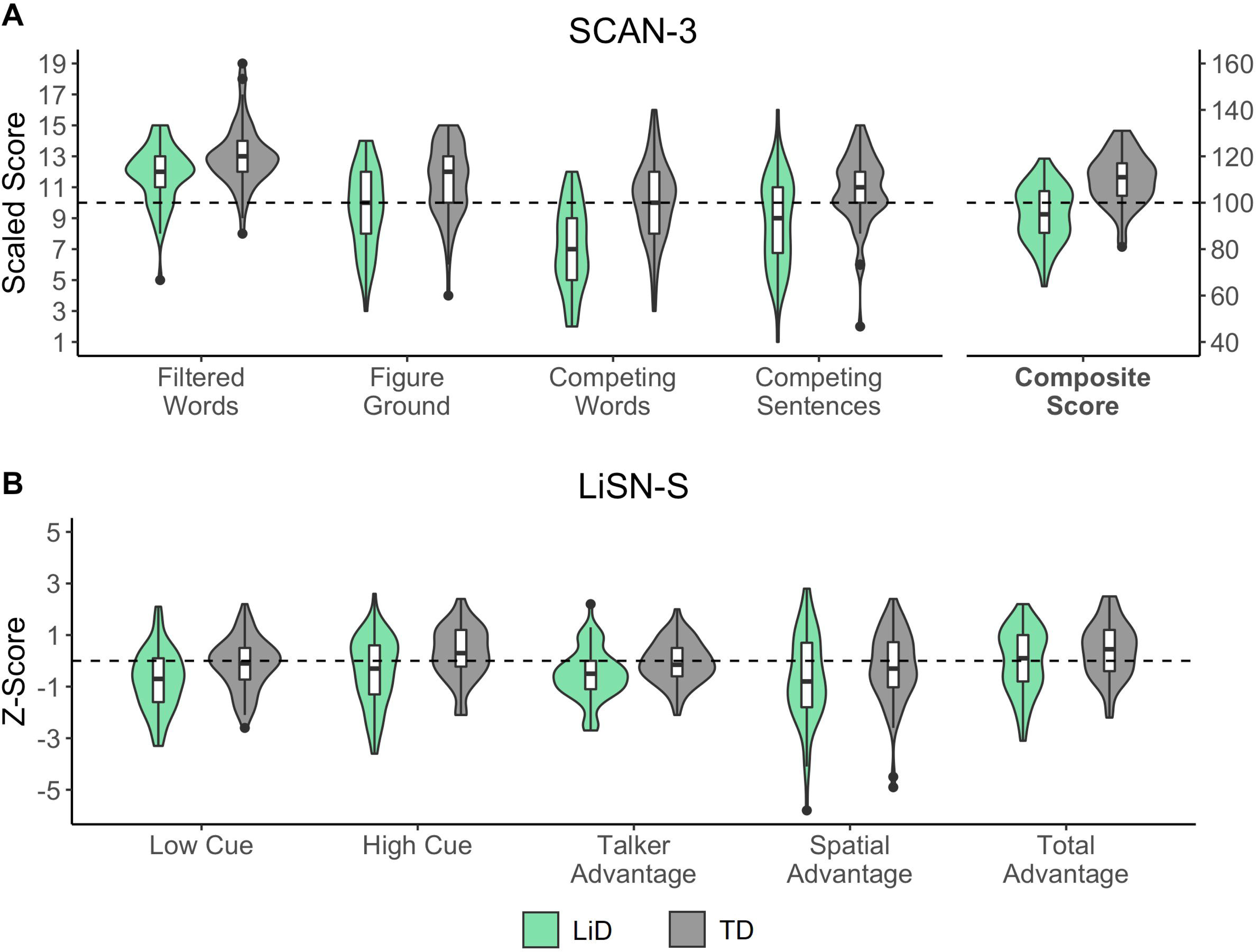
TD children scored more highly than children with LiD on auditory tests. A – SCAN-3:C test for auditory processing disorder in children (Keith, 2009). Violin plots of scores on all subtests and the aggregate (Composite) score. B – LiSN-S listening in spatialized noise test (Cameron & Dillon, 2007). Cue and Advantage scores.

**Figure 3:**
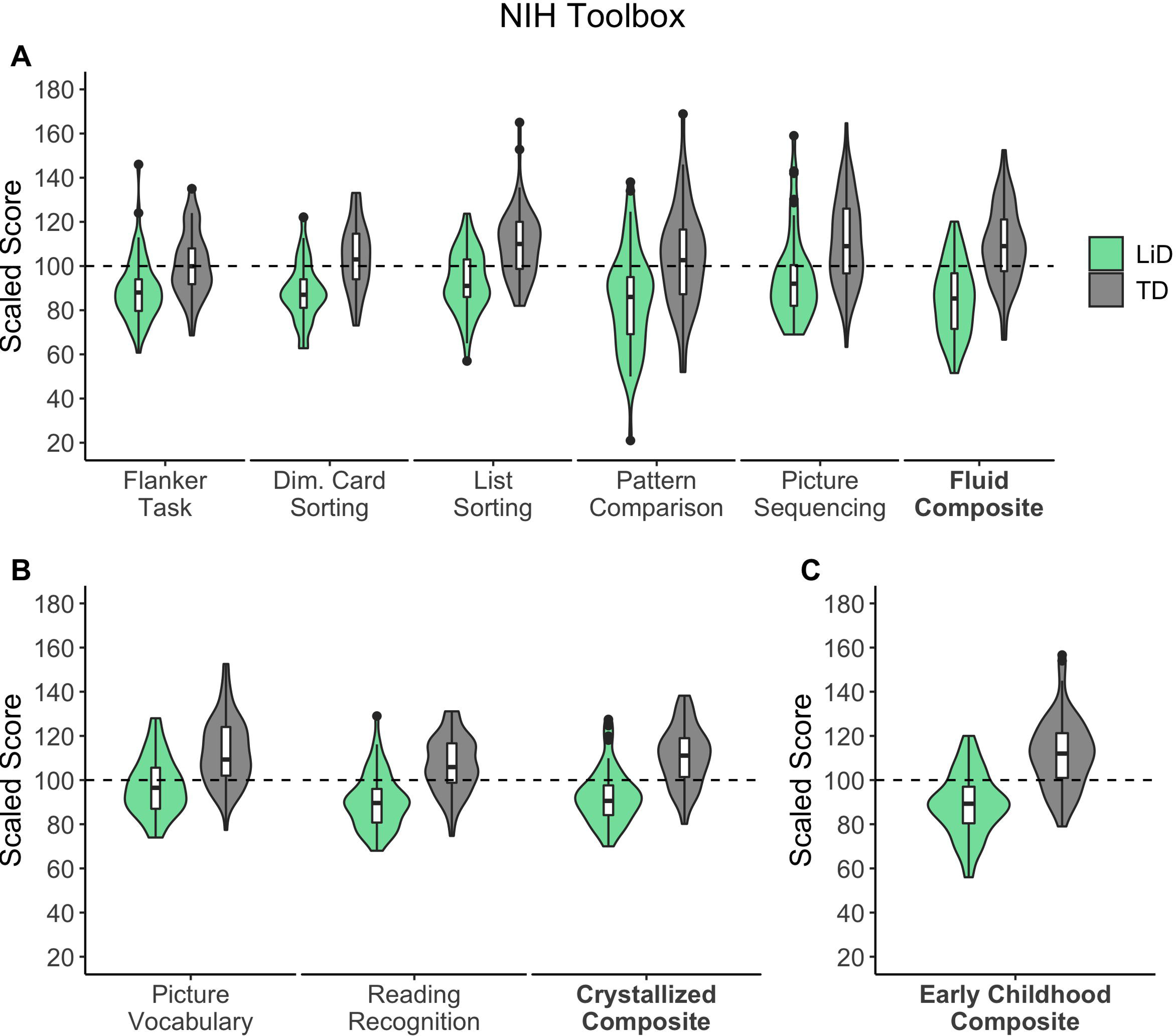
TD children scored more highly than children with LiD on cognitive tests. Scaled scores from the core NIH Cognition Toolbox (Weintraub *et al*., 2013) tests by group. A – individual tests of the Fluid Composite scale. B – individual tests of the Crystallized Composite scale. C – the Early Childhood Composite.

As can be seen in Table 3 and Figure 2B, both the Low Cue [*t*(138) = 3.32, *p* = 0.004, *d* = 0.56] and Talker Advantage [*t*(138) = 3.22, *p* = 0.006, *d* = 0.55] scores were significantly lower for children with listening difficulties than the TD group. No significant differences were observed for the Spatial Advantage score [*t*(138) = -1.64, *p* = 0.41, *d* = 0.28]. Note that Spatial Advantage had more variance, particularly on the lower scoring side, than the other measures. Mean LiSN-S subscore and Advantage z-scores for the TD group were all close to zero, thus matching published US norms (Brown *et al*., 2010). Single-sample *t*-tests confirmed that TD group scores did not differ significantly from those of the normative sample on the LiSN-S Low Cue, Spatial Advantage, or Talker Advantage scores. By contrast, the LiD group means were significantly lower than the normative sample for all three scores (see Supplementary Table 1).

A follow-up analysis of the LiSN-S used a spatial ‘Pattern Score’, developed by Cameron and Dillon (2011) as a quantitative clinical measure of the benefit of adding virtual spatial cues to the information in the Low Cue condition of the LiSN-S (i.e., target and distracting stimuli presented diotically). Pattern Scores did not differ significantly between the LiD (mean = 7.25 dB, n = 69) and TD (mean = 7.29 dB, n = 86) groups [*t*(153) = 0.12, *p* = 1.0, d = 0.02]. However, 7 children in the LiD group and 5 children in the TD group had Pattern Scores within the range to diagnose a ‘spatial processing disorder’ (Cameron & Dillon, 2008; see Discussion).

NIH Cognition Toolbox scores for the LiD group were considerably lower than those for the TD group on the Fluid [*t*(102) = 7.64, *p* < 0.001, *d* = 1.41] and Early Childhood Composite [*t*(102) = 7.64, *p* < 0.001, *d* = 1.50] scores (Table 3, Figure 3). Given the observed differences between the LiD Dx and noDx subgroups on the Crystallized Composite score and the Rey AVL test, these subgroups were separately compared against the TD group. Crystallized Composite scores were considerably lower than the TD group for both the Dx [*t*(78) = 6.08, *p* < 0.001, *d* = 1.66] and noDx [*t*(98) = -6.40, *p* < 0.001, *d* = 1.32] subgroups, thus they were combined in a single LiD group in Figure 3. Similarly profound deficits versus the TD group were observed on the Rey AVL test for both the Dx [*t*(62) = -6.35, *p* < 0.001, *d* = 1.83] and noDx [W = 332, *p* = 0.024, *d* = 0.91] subgroups. Follow-up comparisons of the LiD group (combined across Dx and noDx subgroups) versus the TD group for all of the individual tests contributing to the composite scores were all significant (see Table 3). Comparisons against the normalization samples for the NIH Cognition Toolbox yielded significantly higher scores for the TD group than the samples on many tests, as well as the Fluid, Composite, and Early Childhood composites (Supplementary Table 1). In contrast, the LiD group had significantly lower scores on nearly all tests, except the PSMT and PVT, as well as all three composite scores.

### Four test scores predicted parent-reported listening skills

As introduced in the Methods section, several candidate predictors were explored for inclusion in the regression analysis, including maternal education level, race, SCAN-3:C Composite Score, four LiSN-S scores (Low Cue, High Cue, Talker Advantage, Spatial Advantage), four NIH Cognition Toolbox tests (DCCS, Flanker, PST, PVT), and four interactions between EHF hearing thresholds and measures of listening and vocabulary (LiSN-S Low Cue, Talker Advantage, Spatial Advantage, and NIH PVT). Spearman’s Rho is reported in Table 4 for all candidate predictors that demonstrated univariate association with ECLiPS Total Score with *p* < 0.1. Further statistics including regression coefficients, standard error of the regression coefficient, Student’s *t*-tests, and *p*-values are provided for predictors that were retained in the final model. Notable among these was a specific, significant interaction between the LiSN-S Talker Advantage and EHF threshold.

**Table 4:** Correlations with the ECLiPS Total Score for all continuous predictors that were entered into the stepwise regression. Further statistics yielded from the regression, including regression coefficients and their standard errors, *t*-values, and *p*-values are provided for predictors that were retained in the final model.

|  | **Spearman**<br>**Rho** | **Regression**<br>**Coefficient** | **Standard**<br>**Error** | ***t* value** | ***p* value** |
| --- | --- | --- | --- | --- | --- |
| ***SCAN3:C*** |  |  |  |  |  |
| *Composite Score* | 0.51*** | 0.077 | 0.030 | 2.54 | 0.013 |
| ***LiSN-S*** |  |  |  |  |  |
| *Low Cue* | 0.23* | - | - | - | - |
| *High Cue* | 0.30** | - | - | - | - |
| *Talker Advantage* | 0.28** | 1.06 | 0.372 | 2.84 | 0.006 |
| *Spatial Advantage* | 0.27** | - | - | - | - |
| ***NIH Cognition Toolbox*** |  |  |  |  |  |
| *PVT* | 0.51*** | 0.085 | 0.026 | 3.28 | 0.002 |
| *PSMT* | 0.34*** | - | - | - | - |
| *Flanker* | 0.20 | - | - | - | - |
| *DCCS* | 0.37*** | 0.047 | 0.023 | 2.02 | 0.046 |
| ***Interactions with Extended High Frequency Threshold (EHFT)*** |  |  |  |  |  |
| *EHFT \* LiSN-S Low Cue* | 0.19 | - | - | - | - |
| *EHFT \* LiSN-S Talker Adv.* | 0.27** | - | - | - | - |
| *EHFT \* LiSN-S Spatial Adv.* | 0.17 | - | - | - | - |

\* $p < 0.05$ \*\* $p < 0.01$ \*\*\* $p < 0.001$

One hundred participants (50 LiD, 50 TD) had complete datasets across all predictors. Following stepwise regression, the final model explained 42% of the variance in ECLiPS Total scaled score (F_4,95_ = 17.35, *p* < 0.001). This model had an intercept of -14.15 (SE = 3.10) and included four predictors: the SCAN-3:C composite score, the LiSN-S Talker Advantage score, and the NIH Picture Vocabulary and Dimensional Change Card Sorting scores (Table 4). Figure 4 compares actual ECLiPS Total scaled scores to those predicted by this model, where perfect predictions lie along the diagonal. As expected, the separation between groups was preserved in the predicted scores, though predicted scores tended to be higher than actual scores for the LiD group and lower for the TD group.

**Figure 4:**
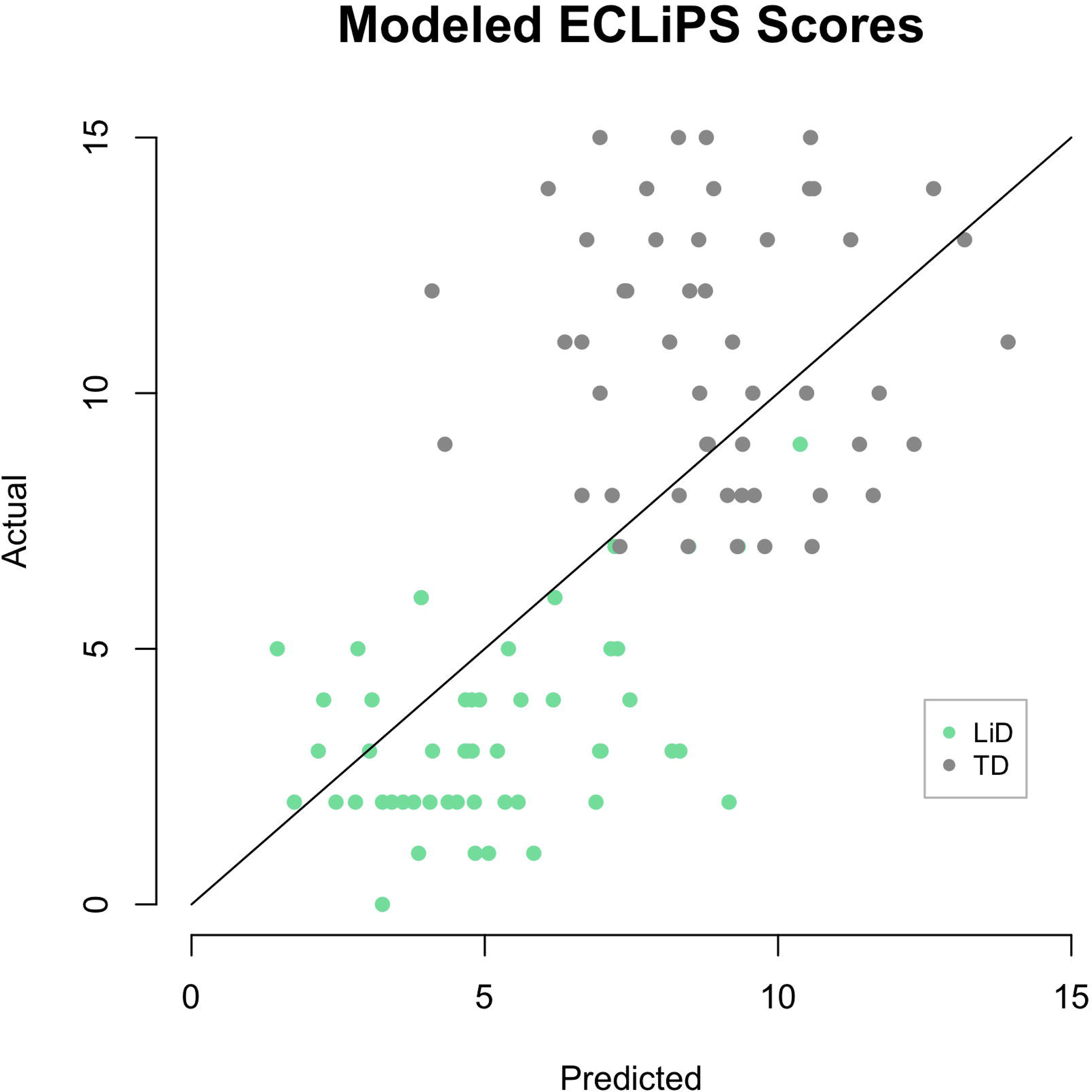
Actual and predicted ECLiPS Total scaled scores. Accuracy of the final equation yielded by stepwise multiple regression, which included four predictors: the SCAN-3:C Composite score, the LiSN-S Talker Advantage score, and the NIH Picture Vocabulary Test (PVT) and Dimensional Change Card Sort test (DCCS) scores. Distance from the diagonal reflects the accuracy of the prediction for each individual. Scores below the diagonal reflect instances in which the predicted scores were higher than actual scores, while those above the line were lower than observed.

### Correlations between measures

In the secondary analysis, pairwise correlations were computed between all variables, treating each as a continuous predictor across groups (i.e., not separated into LiD or TD). These correlations are shown in Tables 5 and 6. The data in Table 5 show that many tests were highly correlated (p < 0.001) with the ECLiPS, SCAN and NIH Toolbox composite measures. In contrast, the LiSN-S Advantage measures correlated poorly with most other, non-LiSN-S measures. A notable exception was a strong correlation between the ECLiPS and the LiSN-S Talker Advantage. The SCAN composite correlated to a highly significant level with the Low and High Cue measures of the LiSN-S, but only weakly and non-significantly with the Spatial and Total Advantage measures. Correlations between LiSN-S and SCAN sub-tests (Table 6), showed that the Filtered Words and dichotic (Competing Words and Competing Sentences) tests, and the Low and High Cue measures accounted for almost all of the relationship between SCAN and LiSN-S. Talker Advantage was marginally related to Competing Sentences and Spatial Advantage to Competing Words. However, when corrected for multiple comparisons (Bonferroni, *p* < 0.01), these relatively weak relationships became non-significant.

**Table 5:**
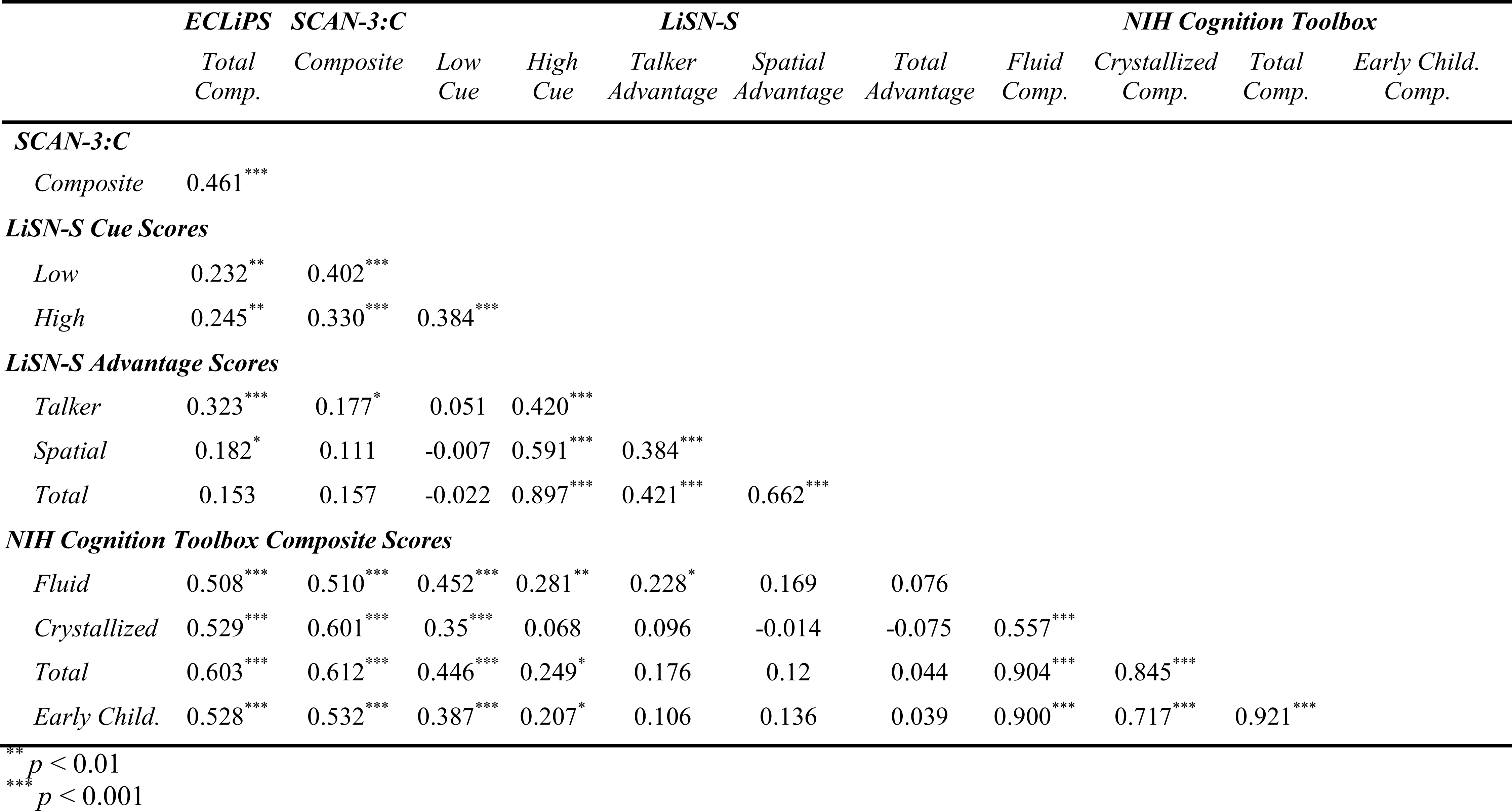
Correlation matrix (Spearman’s Rho) comparing test scores as continuous variables across both LiD and TD children.

**Table 6:** Correlations between subtests of SCAN and LiSN-S for all children (total n = 124) who completed both tests.

|  | *LiSN-S* |  |  |  |  |
| --- | --- | --- | --- | --- | --- |
|  | *Low*<br>*Cue* | *High*<br>*Cue* | *Talker*<br>*Advantage* | *Spatial*<br>*Advantage* | *Total*<br>*Advantage* |
| **SCAN3:C** |  |  |  |  |  |
| *Filtered Words* | 0.293** | 0.187 | 0.040 | -0.050 | 0.058 |
| *Auditory Figure Ground* | 0.097 | 0.128 | 0.114 | 0.069 | 0.108 |
| *Competing Words* | 0.343*** | 0.308*** | 0.172 | 0.212 | 0.159 |
| *Competing Sentences* | 0.360*** | 0.298*** | 0.193 | 0.101 | 0.128 |
| *Composite* | 0.406*** | 0.331*** | 0.177 | 0.111 | 0.156 |

\*\*\* p < 0.01
\*\*\* p < 0.001

## Discussion

In this study, we found that children with LiD, identified primarily on the basis of the ECLiPS caregiver questionnaire, had impaired performance on a broad range of auditory and cognitive tasks relative to age-matched TD children. Tasks included a suite of tests normally used for diagnosing APD, a test for repeating sentences presented against a background of distracting sentences, and a battery of tests measuring fluid and crystallized cognition. Although scores of the TD children were generally higher than those of the normative samples, suggesting the TD children may have been more able than those recruited into other studies, the children with LiD scored almost uniformly below the norm. Conclusions from this study should thus be generalizable.

Given normal peripheral function (Hunter *et al*., 2021), the pattern of deficits observed here with LiD could reflect central auditory (e.g., Cameron *et al*., 2014, 2015; Graydon *et al*., 2017; Moore *et al*., 1991; Pillsbury *et al*., 1991) or general cognitive (e.g., Moore *et al*., 2010; Moore & Dillon, 2018) deficits, or a combination of both. For this reason, the protocol that was developed for our research program cast a wide net, to provide a relatively detailed characterization of the constellation of features associated with this complex clinical construct.

### Diagnosis of APD

A crucial first step in the analysis was to identify any heterogeneities in our sample of children with LiD (as reported on the ECLiPS), some of whom had received clinical diagnoses of APD (Dx), and some of whom had not (noDx). Comparisons between these two subgroups yielded no differences on the ECLiPS Total scaled score, SCAN-3:C composite scaled score, LiSN-S scores (Talker Advantage, Spatial Advantage, and Low Cue scores), or the Fluid and Early Childhood Composite scores of the NIH Cognition Toolbox. However, the Dx subgroup performed significantly more poorly on the NIH Crystallized Composite score and the Rey AVL test. These results suggest that children in the Dx subgroup had more severe problems than those in the noDx subgroup specifically with language.

### Nature of listening difficulties

Comparisons between children with LiD and their TD peers yielded differences across nearly all tested domains of auditory and cognitive function, including the ECLiPS Total scaled score, SCAN-3:C composite scaled score, and all of the NIH Cognition Toolbox composite scores (Fluid, Crystallized, Early Childhood), as well as the Rey AVL test. Several of these tests may tap the same underlying deficit. To disentangle such overlapping contributions, a multivariate regression approach was used. The regression successfully explained 42% of the variance in ECLiPS Total scaled scores using four predictors: the SCAN-3:C composite score, the LiSN-S Talker Advantage score, and the NIH Picture Vocabulary and Dimensional Change Card Sorting scores. These findings suggest that both auditory and cognitive factors make unique, significant contributions to predicting listening skills as captured on the ECLiPS. However, we reason below that just one measure, the LiSN-S Talker Advantage, provided some limited evidence of impaired auditory processing in children with LiD.

### ECLiPS

It is unusual in hearing research for a case-control study such as this to use a subjective assessment, the ECLiPS, as an independent variable. However, that is an accepted clinical practice in the diagnosis of other neurobehavioral disorders, for example developmental language disorder (Bishop & McDonald, 2009) and attention deficit hyperactivity disorder (Pediatrics, 2011). We suggest that a parent or guardian is usually in the best position to judge their child’s behavior; in this case, how their child responds to the challenges inherent in everyday communication. Several aspects of the ECLiPS results assured us that this was a reasonable decision. First, when caregivers responded to our advertisements, there was a high probability that their child’s ECLiPS score would place them into the group identified by the parent (LiD or TD). Second, we were impressed that mean standardized SAP and Listening aggregate scores placed this mid-Western US TD sample closely alongside the English normalization data on which the standard scores were derived (Barry *et al*., 2015). Third, for the children with LiD, the distribution of ECLiPS scores between the various subscales seemed consistent with predominantly receptive speech and language deficits. Whether those deficits are of an auditory, cognitive or mixed origin is not directly addressed by the ECLiPS. Because of the modeling applied to these data, it was possible to observe systematic disparities between caregiver reports and objective measures. To the extent that objective measures capture everyday listening skills, caregivers of children in both groups tended to express a more extreme view of their children’s abilities than the objective data suggested. However, this does not imply that the objective data are more meaningful.

### SCAN-3:C

For many years test batteries such as the SCAN (Keith, 2009) have been used to diagnose APD (American Academy of Audiology, 2010; ASHA, 1996). These batteries test aspects of listening purported to be mediated by central auditory processing, for example ‘closure’, the perceptual capacity to fill in missing or distorted parts of an auditory stimulus, and ‘binaural integration’ (Moncrieff, 2006), the ability to identify different words simultaneously presented to each ear. According to the SCAN manual, closure is represented by the Filtered Words subtest and binaural integration by the Competing Words – Directed Ear subtest. In this study, SCAN composite scores differentiated between the two groups so successfully that the SCAN composite was one of the four measures left in the final multivariate regression that explained nearly half the total sample variance on the ECLiPS. It was thus a strong predictor of LiD.

### LiSN-S

We chose the LiSN-S (Cameron & Dillon, 2007) in this study because it is a multi-faceted test of spatial and speech hearing, and was designed specifically for pediatric assessments. It also features ‘derived’ (also called ‘difference’ or ‘subtraction’) testing that we and others have argued can separate cognitive and sensory contributions to auditory perception when comparing performance within and between listeners (Dillon *et al*., 2014; Moore, 2012; Moore *et al*., 2010; Moore & Dillon, 2018). For the LiSN-S, performance on individual measures, for example the Low Cue and High Cue SRT, reflects both the ability to hear the target sentence against the distracting sentences *and* the ability of the listener to attend selectively to the target, to decode the speech signal, to remember the words in the target sentence, and to repeat each sentence orally. In contrast, for the three derived Advantage measures (Talker, Spatial and Total), performance reflects only the ability to hear the target sentence, assuming that the attention, decoding, memory, linguistic, and reproduction aspects of the task are identical for each of the two contributing individual measures from which each Advantage measure is derived. This assumption appears to be only partly fulfilled for the LiSN-S. The Spatial Advantage relies only on the spatial separation of the distracting talkers, a physical acoustic manipulation. However, the Talker Advantage employs different individual talkers as the distractors, introducing a decoding, a linguistic and, possibly, an attention difference, as well as an acoustic difference between the underlying test conditions.

While Low and High Cue SRTs were higher (i.e., poorer) in the LiD than in the TD group, among the Advantage scores, only the Talker Advantage was significantly poorer in the LiD group, while the Spatial Advantage showed a non-significant trend in that direction. These results are surprising in two respects. The poorer Talker Advantage represents the first and only example of which we are aware of a group difference in this measure in any study (e.g., Cameron & Dillon, 2008). Furthermore, despite some possible cognitive contributions to this metric, the finding of a significant deficit of the LiD group on a derived measure of hearing may still be direct evidence for impaired auditory system function. This possibility is considered further below in the context of correlations between LiSN-S thresholds, SCAN and EHF measures.

It has been suggested that deficits in the LiSN-S spatial measures are the basis for a specific and treatable disorder termed spatial processing disorder (SPD; Cameron *et al*., 2011, 2012, 2014; Cameron & Dillon, 2008) that is diagnosed using the LiSN-S Pattern Score. Here, we found evidence for SPD among both LiD and TD groups of children. However, neither the proportion of children with SPD nor the Pattern Score differed significantly between groups, suggesting that SPD is not predictive, and may not be representative, of the listening problems identified by the ECLiPS.

### NIH Cognition Toolbox

One of the most consistent and largest deficits experienced by the children with LiD in this study was broadly-specified cognitive function, as also recognized in other recent studies (Moore *et al*., 2010; Seeto *et al*., 2021; Tomlin *et al*., 2015). Scores on individual tests were highly correlated and uniformly reduced relative to TD children, suggesting that cognitive function is a major contributor to LiD. Both the Picture Vocabulary and Dimensional Change Card Sorting tests contributed to the final regression model, so further discussion will focus on them. The PVT is an index of language and is highly associated with crystallized intelligence and success in school and work. The DCCS, on the other hand, indexes attention switching, which is an aspect of executive function (Weintraub *et al*., 2013a). Scores on the PVT and DCCS were highly significantly correlated, despite their attribution to different subdomains of cognition and correspondingly different neural pathways. Further studies in our laboratory are investigating the functional neuroanatomy of LiD in this same sample of children using MRI. However, the finding of language and executive function deficits suggests a LiD profile that extends well beyond any notion of a specific auditory processing disorder.

#### Auditory or cognitive?

Although the SCAN correlated highly with the ECLiPS, a key question remains whether the SCAN tests are primarily markers of central auditory or of cognitive function (Moore, 2018). Correlations between the SCAN and the cognitive and LiSN-S variables provide insight into this question. The SCAN total scaled score correlated significantly with all of the domain composites of the NIH Cognition Toolbox and both the Low Cue and High Cue individual scores of the LiSN-S. Critically, the SCAN correlated less well with the derived scores of the LiSN-S (Spatial, Talker, Total Advantage). Similarly, the Low Cue score of the LiSN-S correlated well with tests of cognition, as above, but the derived scores of the LiSN-S notably lacked such correlations.

Because the derived scores of the LiSN-S reflect a more sensory measure of auditory perception, these data suggest overall that the SCAN is more sensitive to cognitive than to auditory factors that contribute to perception. Although the LiSN-S Talker Advantage was significantly correlated with the SCAN Composite, we suggest above that this could be due to remaining cognitive contributions to the Talker Advantage. The inclusion of the SCAN in the final regression model, and the strong correlation between the ECLiPS Total scaled score and the SCAN Composite suggest the possibility that one or more of the SCAN subtests may be valid predictors of a truly auditory processing disorder. However, as shown in Table 6, even the relation between SCAN dichotic subtests and LiSN-S Advantage measures is weak relative to that between SCAN dichotic subtests and LiSN-S Cue measures.

A final point concerning LiSN-S Talker Advantage is that it, alone among the LiSN-S measures, interacted significantly with EHF threshold. Recently, we have shown that EHF hearing plays a role in speech-in-noise perception in adults (Hunter *et al*., 2020; Motlagh Zadeh *et al*., 2019). Recurrent otitis media in childhood, especially that treated with tympanostomy tubes, appears to contribute to reduced EHF thresholds, as shown in both groups of the sample examined here (Hunter *et al*., 2021) and in other studies (Hunter *et al*., 1996). The robust relationship between Talker Advantage and ECLiPS scores may thus receive a contribution from a truly auditory process, EHF hearing, the sensitivity of which is presumably determined by ear function. However, these influences are likely to be minor compared with the dominance of cognitive function in listening.

Overall, the results reported here demonstrate a major influence of impaired cognitive function on listening difficulties of children with normal audiograms, and provide some support for an additional auditory contribution of subclinical hearing loss. Evaluation of children with complaints of listening difficulties should include standardized caregiver observations, EHF audiometry, and consideration of broad cognitive abilities, beyond auditory tests.

## Data Availability

The data described in this manuscript is not available for open access.

## Acknowledgments

This research was supported by grant DC014078 from the National Institute of Deafness and other Communication Disorders, and by the Cincinnati Children’s Research Foundation. DRM was supported in part by the NIHR Manchester Biomedical Research Centre. Thanks to Allison Bradley for her contributions to the project.

## Contributions of authors to research

L.P. analyzed and interpreted data, and co-wrote the paper; L.H. contributed to the design of the study, provided input to the analysis, and was involved in critical revisions; L.M.Z. collected, analyzed, and interpreted data, contributed to the writing, and was involved in critical revisions; H.J.S. collected, analyzed, and interpreted data, and was involved in critical revisions; N.T.S. collected, analyzed, and interpreted data, contributed to the writing, and was involved in critical revisions; A.P. collected, analyzed, and interpreted data, contributed to the writing, and was involved in critical revisions; L.L. analyzed and interpreted data and was involved in critical revisions; D.R.M. designed the study, analyzed and interpreted data, and co-wrote the paper.

---

**Supplementary Table 1:** Single-sample *t*-tests comparing LiD and TD groups separately against the normative samples for each standardized test. All *p*-values are adjusted for multiple comparisons. Effect sizes are expressed using Cohen’s d.

|  | TD Group |  |  |  | LiD Group |  |  |  |
| --- | --- | --- | --- | --- | --- | --- | --- | --- |
| | $t$ | df | $p$ | $d$ | $t$ | df | $p$ | $d$ |
| ***ECLiPS Composite Scores*** |  |  |  |  |  |  |  |  |
| *Total* | 2.98 | 78 | 0.004 | 0.34 | -32.73 | 66 | < 0.001 | 4.00 |
| ***CCC-2 Composite Scores*** |  |  |  |  |  |  |  |  |
| *GCC* | 12.14 | 78 | < 0.001 | 1.37 | -10.64 | 63 | < 0.001 | 1.33 |
| ***SCAN-3:C*** |  |  |  |  |  |  |  |  |
| *Comp.* | 7.64 | 68 | < 0.001 | 0.92 | -3.24 | 58 | 0.002 | 0.42 |
| ***LiSN-S Cue Scores*** |  |  |  |  |  |  |  |  |
| *Low* | -1.14 | 75 | 0.78 | 0.17 | -5.03 | 63 | < 0.001 | 0.63 |
| ***LiSN-S Advantage Scores*** |  |  |  |  |  |  |  |  |
| *Spatial* | -1.47 | 75 | 0.44 | 0.13 | -3.08 | 63 | 0.009 | 0.38 |
| *Talker* | -0.56 | 75 | 1.00 | 0.06 | -4.38 | 63 | < 0.001 | 0.55 |
| ***NIH Cognition Toolbox Tests*** |  |  |  |  |  |  |  |  |
| *Flanker* | 0.32 | 56 | 1.00 | 0.04 | -5.46 | 56 | < 0.001 | 0.72 |
| *DCCS* | 2.09 | 56 | 0.45 | 0.28 | -6.65 | 56 | < 0.001 | 0.88 |
| *LSWM* | 5.20 | 66 | < 0.001 | 0.63 | -3.81 | 55 | 0.004 | 0.51 |
| *PCPS* | 1.07 | 51 | 1.00 | 0.15 | -5.00 | 53 | < 0.001 | 0.68 |
| *PSMT* | 4.66 | 72 | < 0.001 | 0.55 | -1.99 | 51 | 0.58 | 0.28 |
| *PVT* | 7.27 | 71 | < 0.001 | 0.86 | -1.56 | 58 | 1.00 | 0.20 |
| *RR* | 4.34 | 63 | < 0.001 | 0.54 | -6.29 | 54 | < 0.001 | 0.85 |
| *AVL* | 2.95 | 47 | 0.054 | 0.43 | -4.50 | 40 | < 0.001 | 0.70 |
| ***NIH Cognition Toolbox Composite Scores*** |  |  |  |  |  |  |  |  |
| *Fluid* | 3.69 | 50 | 0.006 | 0.52 | -6.10 | 44 | < 0.001 | 0.91 |
| *Crystallized* | 6.42 | 62 | < 0.001 | 0.81 | -4.52 | 53 | < 0.001 | 0.61 |
| *Early Childhood* | 5.57 | 55 | < 0.001 | 0.74 | -5.34 | 47 | < 0.001 | 0.77 |

